# Deciphering the heterogeneous niche in the tumor progression of hepatocellular carcinoma: a Spatial single-cell landscape and multi-omics atlas analysis

**DOI:** 10.1101/2022.03.09.22272063

**Authors:** Jiazhou Ye, Yan Lin, Zhiling Liao, Xing Gao, Cheng Lu, Lu Lu, Julu Huang, Xi Huang, Tao Bai, Jie Chen, Xiaobo Wang, Min Luo, Mingzhi Xie, Feixiang Wu, Guobin Wu, Liang Ma, Bangde Xiang, Yongqiang Li, Hongping Yu, Xiaoling Luo, Rong Liang

## Abstract

Hepatocellular carcinoma (HCC) is an invasive disease which is characteristic with highly heterogeneous molecular phenotype, rich blood supply, and unique immune niche, therefore it is of great significance to explore the tumor heterogeneous niche and clonal evolution progress of these malignant cells. Based on the advance in single-cell technology, spatial transcriptome technology, and Oxford nanopore technology, this study innovatively reconstructed and delineated the heterogeneity of the HCC tumor niche and its tumor progression pattern. Our results showed that the copy number variation (CNV) of cells in cancer lesions and liver cirrhosis lesions of the same patient is basically the same and is mainly regulated by transcription factors such as TP53, HOXA7, FOXN3, and PPARG, suggests that malignant cells of common origin gradually evolve into different lesions in a very rare numbers of different CNVs, which are mainly regulated by expression patterns and mediate the heterogeneity between the tumor and cirrhosis lesions. Angiogenesis-related genes (SREBF1, ZNF585A, and HOXB5) may mediate communication between HCC subpopulations and endothelial cells via exosomes, thereby contributing to the angiogenic niche before HCC metastasis. In addition, numerous CNVs were found in patients with early recurrent HCC, and these mutated genes is the potential niche genes for the early tumor recurrence. In summary, this study provides a general transcriptional landscape of the ecological structure of HCC, systematically maps the molecular, cellular, and spatial composition of different HCC cell niches, and provides a scientific and theoretical basis at the molecular and cellular levels for personalized and accurate treatment strategies for HCC.

## Introduction

Hepatocellular carcinoma (HCC) is the sixth most common malignancy and the second leading causes of cancer-related deaths worldwide, which is characteristic with high aggressiveness and invasiveness ^[1, 2]^. Although progress has been made greatly in regional therapies in the past few decades, the prognosis of patients remains poor. So far, the five-year recurrence rate for radical HCC treatments (surgical resection or radiofrequency ablation) still beyond 70% ^[3, 4]^, even in patients with early HCC (<3 cm) who underwent surgery, the five year survival rate was unsatisfactory (47–53%) ^[5, 6]^. Besides, HCC is still highly resistant to systematic anti-cancer therapies owing to its highly heterogeneous molecular phenotype, rich blood supply, and unique immune niche. Conventional chemotherapy, small molecular targeted therapy and immunotherapy all provide limited survival benefit ^[7-12]^.

Intratumoral heterogeneity is one of the main reasons why current therapies are ineffective for most types of cancers, especially HCC ^[13]^. However, most current theories describing the generation of intratumor heterogeneity are based on tumor evolution ^[14]^. In tumor cells, subclones arise when novel genomic aberrations lead to distinct cell subpopulations, which are driver mutations in key oncogenes or tumor suppressor genes that confer an adaptive advantage to tumor cells ^[15]^. For example, tumor cells can develop drug resistance through gene amplification of therapeutic targets, point mutations that affect the ability of the therapy to inhibit oncogenic pathways, and/or amplification/suppression of other genes that compensate for drug-suppressed oncogenes ^[16]^. In addition, tumor metastasis and recurrence contributes to the weakness of locally targeted therapies. In recent years, molecules and cells identified in distant metastatic tissues of different tumor animal models, including primary tumor-derived soluble factors, vesicles, exosomes, and bone marrow derived cells (BMDCs), have gradually confirmed the existence of a premetastatic niche ^[17, 18]^. The premetastatic niche refers to a microenvironment that is well prepared for tumor cell colonization and the spread to distant organ sites ^[19, 20]^. Signals from the primary tumor mobilize and adapt immune cells and communicate directly with distant niche cells to induce broad adaptations in target organs, including the induction of angiogenesis, inflammation, extracellular matrix remodeling, and metabolic reprogramming, which interact together to promote the formation of pre-transfer niches ^[21]^. HCC metastasis to specific organs is associated with the generation of pre-metastatic niches ^[22, 23]^; however, the exact mechanism remains unclear. Therefore, a comprehensive and accurate understanding of the genomic structure of HCC, including mutations, copy number variation (CNV), RNA expression, tumor immune microenvironment landscape, and clarifying the tumor heterogeneous niche and its clonal evolution model in the HCC tumor microenvironment are very important for the development of personalized therapy.

Single-cell analysis, which enables us to better define tumor cell populations and identify potential targets for immunotherapy or combination therapy, has been widely used in HCC research ^[24]^. In addition, spatial omics can reveal the impact of cell spatial distribution on a disease by studying the relative positional relationship of cells in tissue samples ^[25]^. Therefore, based on increasingly mature single-cell technology, spatial transcriptome (ST) technology, and Oxford nanopore technology (ONT), we innovatively reconstructed and delineated the heterogeneity of HCC tumor niches and their clonal evolution patterns.

## Methods

### Human samples

Six HCC patients and one patient with hemangioma included in this study agreed to a diverse library and sequencing protocol covering all research procedures, which was approved by the ethics review committee of the Guangxi Medical University Cancer Hospital. Demographic and clinical data are presented in Table S1. According to the research needs, six tumor tissues and one liver cirrhosis nodule were obtained from HCC patients as a case, and one normal liver tissue was obtained from a hemangioma patient as a control for sequencing. Sequencing was performed using BMKCloud (Biomarker Technologies Corporation, Beijing, China).

### Spatial transcriptome sequencing

Frozen embedded tissue stored at –80°C was sliced to extract RNA and subjected to quality inspection. The RNA integrity number (RIN) value was required to be ≥7. Then, the tissue permeabilization time was explored for the sample, and tissue permeabilization and mRNA reverse transcription were performed to synthesize cDNA according to the permeabilization time determined by tissue optimization. Next, synthesis and denaturation of the second strand of cDNA was performed. The cycle number of cDNA amplification was determined by quantitative polymerase chain reaction (qPCR), and the cDNA was purified and quality controlled, after which the gene expression library was constructed and quality checked. After the library passed quality inspection, the Visium spatial gene expression library was sequenced using the Illumina NovaSeq 6000 platform, and the sequencing strategy used was PE150.

### Single-cell sequencing

Sample preparation and cDNA library construction were performed as described in the 10X Genomics Single Cell 3’ Kit v3.1 User Guide. Droplets of latex gel beads (GEMs) containing cells were obtained using microfluidic technology. The GEMs were then broken, and the cDNA was recovered and amplified by PCR to complete the construction of the cDNA library. The cDNA product and library concentration were detected using a Qubit 4.0, and a Qseq400 bioanalyzer was used to detect the insert size of the cDNA library to ensure a qualified insert size, single peak shape, no spurious peaks, no adapters, and no primer dimers. Finally, the sample library was sequenced using a Novaseq 6000 instrument on an Illumina platform. After identifying the Casava bases, the obtained raw image files were converted into sequence files and stored in the FASTQ format. The sequencing data were compared and quantified using CellRanger, the official 10X Genomics software.

### The construction of single-cell maps and annotation of cell types

For single-cell sequencing (scRNA-seq) data, the IntegrateData function in the Seurat package ^[26]^ was used to merge single-cell data and cell cluster analysis was performed according to the default parameters. The clustering results were uniformly reduced and visualized using the uniform manifold approximation and projection for dimensionality reduction (UMAP) algorithm^[27]^, projected onto a 2-dimensional image, defined as a single-cell atlas. In addition, the FindAllMarkers function of the Seurat package was used to identify the markers for each cell cluster. Referring to the SingleR package^[28]^, the defined cell types were annotated into the single-cell atlas according to the known cell markers.

### CNV estimation in single cells

Single-cell CNV estimates were performed using the copy number karyotyping of Aneuploid Tumors (CopyKAT) method^[29]^. CopyKAT calculates the genome copy number distribution of a single cell by combining Bayesian methods and hierarchical clustering and defines the subclone structure.

### Functional enrichment analysis

To further explore the biological processes and pathways involved in the markers of each cell cluster, the R software package clusterProfiler^[30]^ was used to perform enrichment analysis on the biological processes (BP) and KEGG signaling pathways of Gene Ontology (GO) for the markers, P value < 0.05 for BP and KEGG signaling pathways were considered significant.

### Pseudotime and RNA rate calculation

Pseudotime explores the cellular trajectories of cells transitioning from one state to another during development, disease, and throughout life, based on changes in gene expression in different cell subsets over time. In this study, the R language package Monocle 3 ^[31]^ was used to reconstruct the differentiation and developmental trajectory of the single-cell atlas of HCC patient tumor cells and to simulate the evolutionary trajectory of malignant cancer cell subclones.

In addition, RNA rates (the time derivative of the gene expression state) can be used to predict the future state and ultimate fate of individual cells, thereby dissecting their developmental lineage and cellular dynamics. Here, referring to velocyto.R ^[32]^ by Gioele La Manno, RNA rates were directly calculated based on distinguishing between unspliced and spliced mRNAs in single-cell maps.

### Gene regulatory network

Single-cell regulatory network inference and clustering (SCENIC) is a tool used to infer gene regulatory networks based on single-cell expression profiles to identify cell states ^[33]^. In this study, we used the Python module tool pySCENIC ^[34]^ to analyze and comprehensively reconstruct gene regulatory network (GRN) centered on transcription factors. The workflow starts with a count matrix describing the gene abundance of all cells and consists of three stages. First, co-expression modules were inferred using the per-target regression method (GRNBoost2). Indirect targets were pruned from these modules using cis-regulatory motif discovery (cisTarget). Finally, the activity of these regulons was quantified using the enrichment fraction of the regulon target genes (AUCell). Based on the cellular activity patterns of these regulons, nonlinear projection methods can be used to display the visual groupings of cells.

### Data Analysis and Statistics

All bioinformatic analyses in this study were performed using the Bioinforcloud platform (http://www.bioinforcloud.org.cn).

## Results

### Global single-cell ecology and spatial distribution of HCC

According to the UMAP method, 54 cell clusters, including HCC malignant cells, myeloid cells, and lymphocytes, belong to the common cell population in HCC (Figure 1A). Further CNV estimation using CopyKAT in patients with HCC allowed for the identification of aneuploid tumor cells in the single-cell transcriptome data of patients with HCC (Figure 1B). Cancer cells in solid tumors typically contain aneuploid copy number events in their genomes, whereas most stromal and immune cells have diploid copy number profiles. These cell subpopulations expressed molecular markers consistent with previously known laboratory markers (Figure 1C). Mapping patients using single-cell profiles indicated that the data were free from noise contamination, such as batch effects (Figure 1D, E). The cell type ratio analysis reflected the widespread presence of HCC malignant cells, CD8+ T cells, B cells, and Kupffer cells (hepatic macrophages) in patients with HCC (Figure 1F). Interestingly, the widespread presence of these cells was also observed in the ST analysis (Figure 1G). In patients, ST analysis was performed on HCC malignant cells and tumor-associated spot clusters were mapped to HCC malignant cells using scRNA-seq (Figure 1H). Taken together, these data provide a representative cellular atlas of HCC.

**Figure 1:**
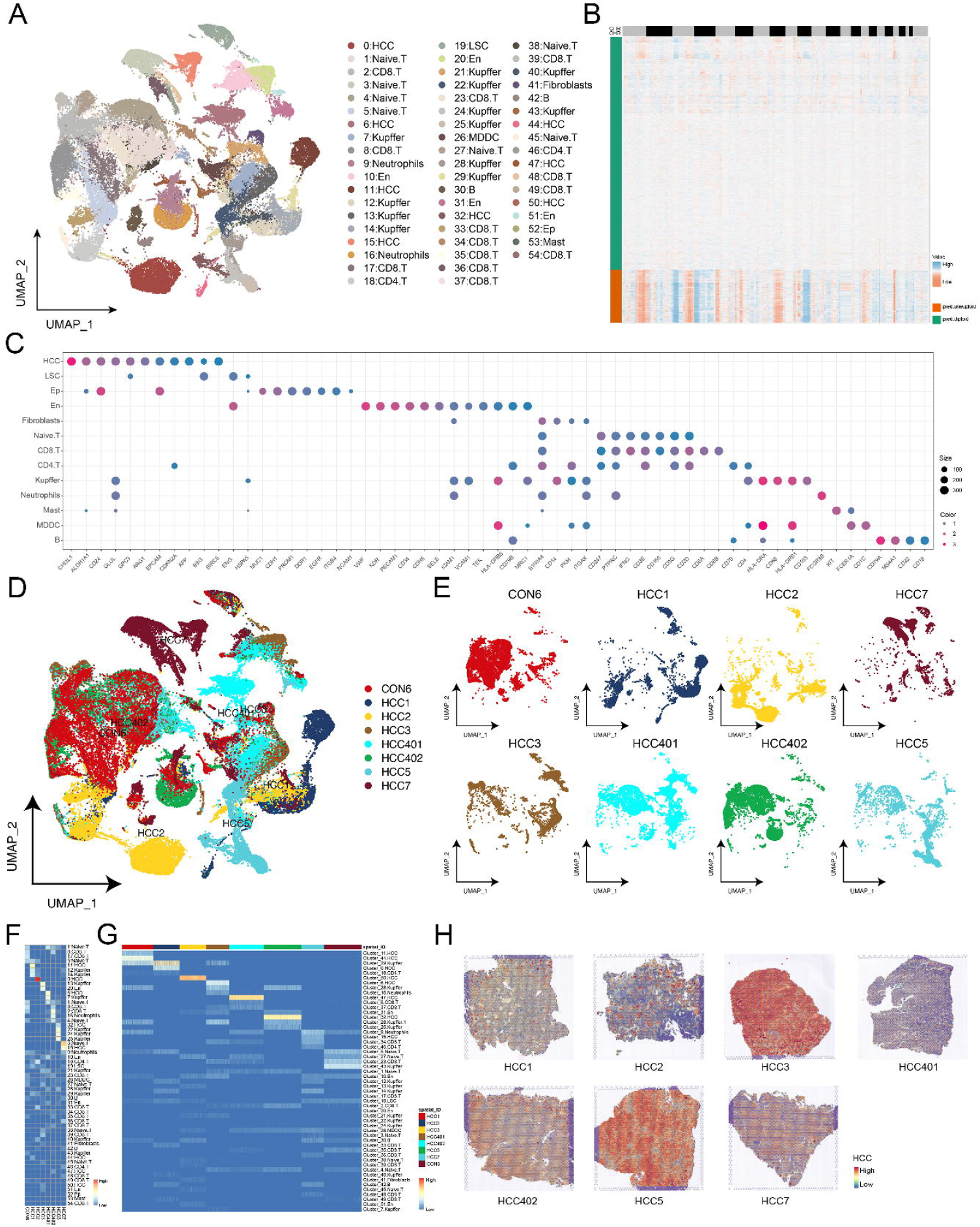
Single-cell and spatial transcriptomes reveal the global single-cell ecology and spatial distribution of hepatocellular carcinoma. A. The single-cell atlas showing that the cells of all samples are divided into different cell subsets. B. Chromosome heatmap showing single-cell profiles of liver cancer patients based on copy number variation (CNV) estimation. C. Expression of marker genes annotating cell types. D–E. Single-cell atlas map single cells from patient and control donors. F. Heatmap mapping the proportion of cell types in each sample in a single cell. G. Heatmap maps the proportion of cell types in spatial resolution. H. Hematoxylin and eosin (H&E) staining of patient sections and unbiased clustering of spots based on HCC malignant cell gene expression within individual spots.

### HCC heterogeneity mainly stems from the diversity of tumor cells

Single-cell analysis revealed the existence of several different subpopulations of HCC (Figure 2A), which may contribute to the heterogeneity of HCC. The expression of markers guiding the annotation of each HCC subpopulation was demonstrated at both the single cell (Figure 2B) and spatial transcriptome (Figure 2C) levels, and the markers were all specifically expressed in the HCC subpopulation, indicating that these markers are of guiding significance. Subsequent enrichment analysis of the HCC subpopulations revealed that the markers were all involved in drug metabolism and other related pathways, mainly cytochrome P450 (Figure 2D). These markers of subpopulations are differentially expressed and significantly involved in different signaling pathways (P value < 0.05) and are one of the main reasons for the heterogeneity of HCC.

**Figure 2:**
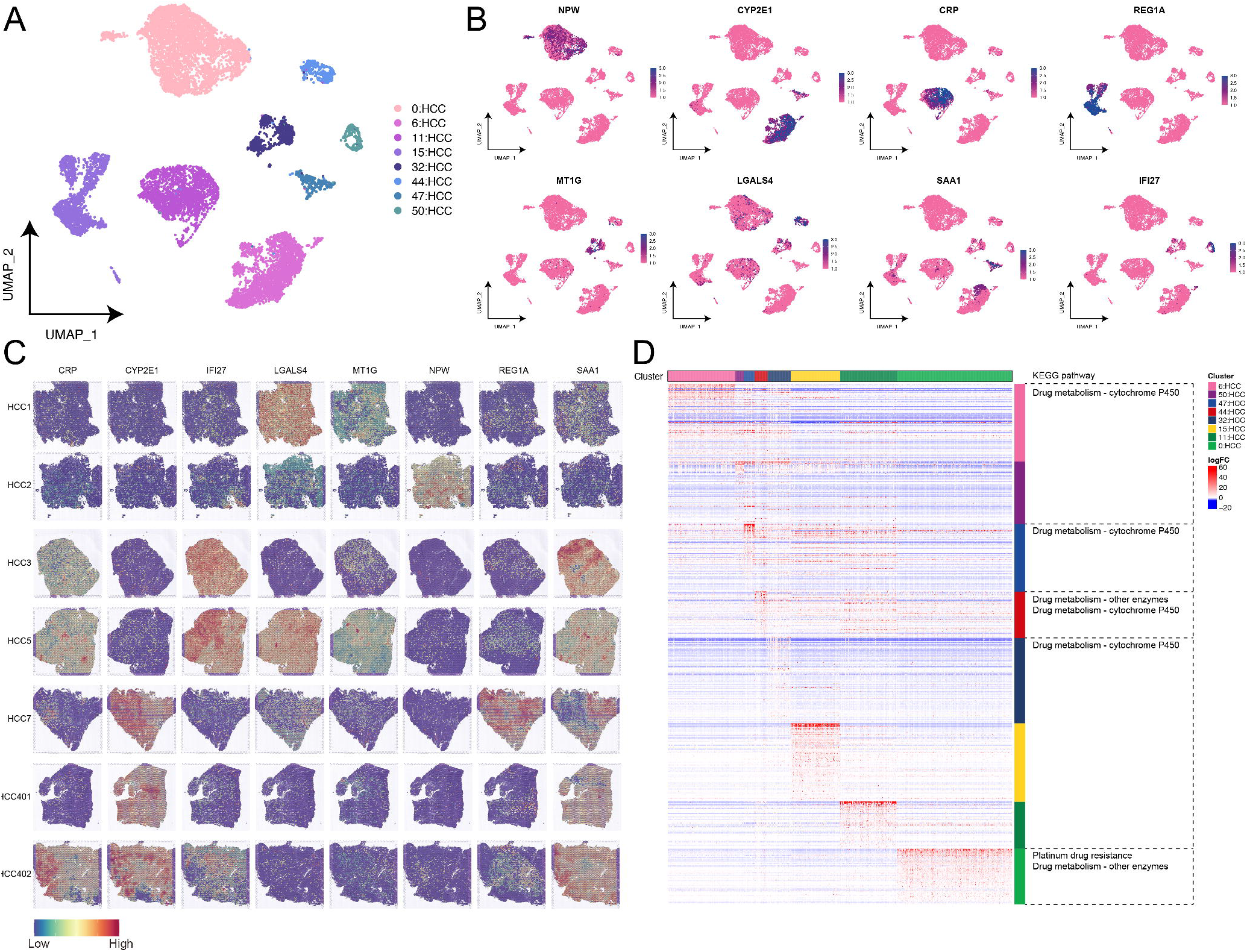
Subpopulation Analysis of hepatocellular carcinoma cells. A. The single-cell profiling maps subpopulations of HCC. B. Single-cell atlas maps specific markers of HCC subpopulations. C. Spatial transcriptome mapping maps specific markers of HCC subpopulations. D. The heatmap-pathways showing the biological functions and signaling pathways in which the markers of HCC subgroups are significantly involved (P value <0.05).

### Angiogenesis in HCC

HCC is characterized by a rich blood supply, and endothelial cells are crucial in HCC angiogenesis. Studying the characteristics of endothelial cells is important to reveal differences in the rate of HCC progression and sensitivity to targeted therapies. A large number of endothelial cells were found in all HCC subpopulations (Figure 3A), and the specific expression of markers guiding the annotation of endothelial cells in endothelial cell subpopulations was demonstrated (Figure 3B). Functional enrichment analysis was performed to explore the biological functions of the HCC and endothelial cell subpopulations. The results revealed that the HCC subpopulation was mostly involved in tumor-related pathways, such as the PI3K-AKT and HIF1 signaling pathways, while resistance to epidermal growth factor receptor tyrosine kinase inhibitors was significantly activated in endothelial cells in HCC (Figure 3C). A gene regulatory network with transcription factors such as fulcrums was subsequently explored, which was organized into seven modules (Figure 3D), with SREBF1, ZNF585A, and HOXB5, which are transcription factors that regulate endothelial cells in HCC patients. In addition, the expression of exosome marker genes (CD9, CD63, and CD81) was observed in both HCC subpopulations and endothelial cells (Figure 3E), suggesting that HCC subpopulations and endothelial cells achieve intercellular communication through exosomes, and thus, participate in the formation of angiogenic niches in HCC.

**Figure 3:**
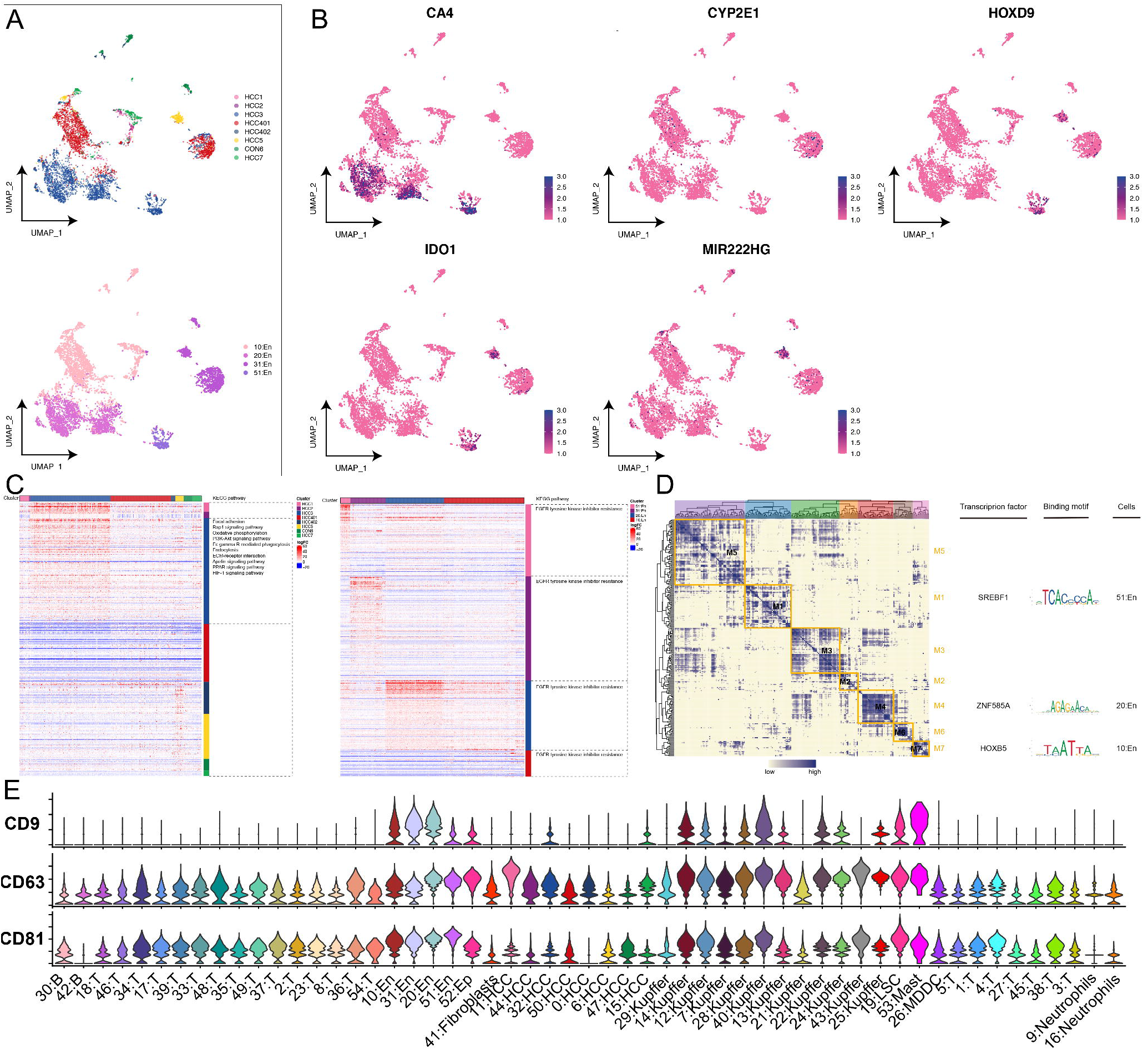
Exploration of the angiogenic niche in hepatocellular carcinoma (HCC). A. The single-cell atlas maps the single-cell landscape of a patient exhibiting liver cancer endothelial cells. B. The single-cell atlas maps specific markers of endothelial cell subsets in patients with angiogenesis (HCC3, HCC401, and HCC5). C. The heatmap pathways showing the endothelial cell subsets and the markers in patients with angiogenesis and their significantly involved biological functions and pathways (P value <0.05). D. The modular heatmap transcription factor (motif-Logo) subpopulation showing the gene regulatory network of angiogenic subpopulations of HCC. E. The violin plot showing the expression of exosome marker genes in HCC and epithelial cells.

### Specific immune cells are closely related to tumor immunity

The development of HCC is also closely associated with immune cells, and after reaggregation of CD8T+ cells, 13 major CD8T+ cell subpopulations were observed, which, interestingly, had notable patient heterogeneity in their abundance (Figure 4A). Some subpopulations expressed representative immune-related genes, including IFNG, GNLY, and CCL20 (Figure 4B). Similarly, re-clustering of B cells revealed two subpopulations of B cells (Figure 4C), which also showed patient heterogeneity. We also observed high expression of CD81, HLA-DRA, and IGHG1 in different B cell subpopulations (Figure 4D). In addition, we found that Kupffer cells were widespread in HCC patients; therefore, re-clustering of Kupffer cells captured twelve cell subpopulations (Figure 4E), and each subpopulation was patient-heterogeneous. Remarkably high expression levels of CCL2, IL1B, and MARCO were also observed in the Kupffer cell subpopulation (Figure 4F). After re-clustering the different immune cells, they were all found to be patient-heterogeneous, suggesting that the development of HCC is closely related to immunity.

**Figure 4:**
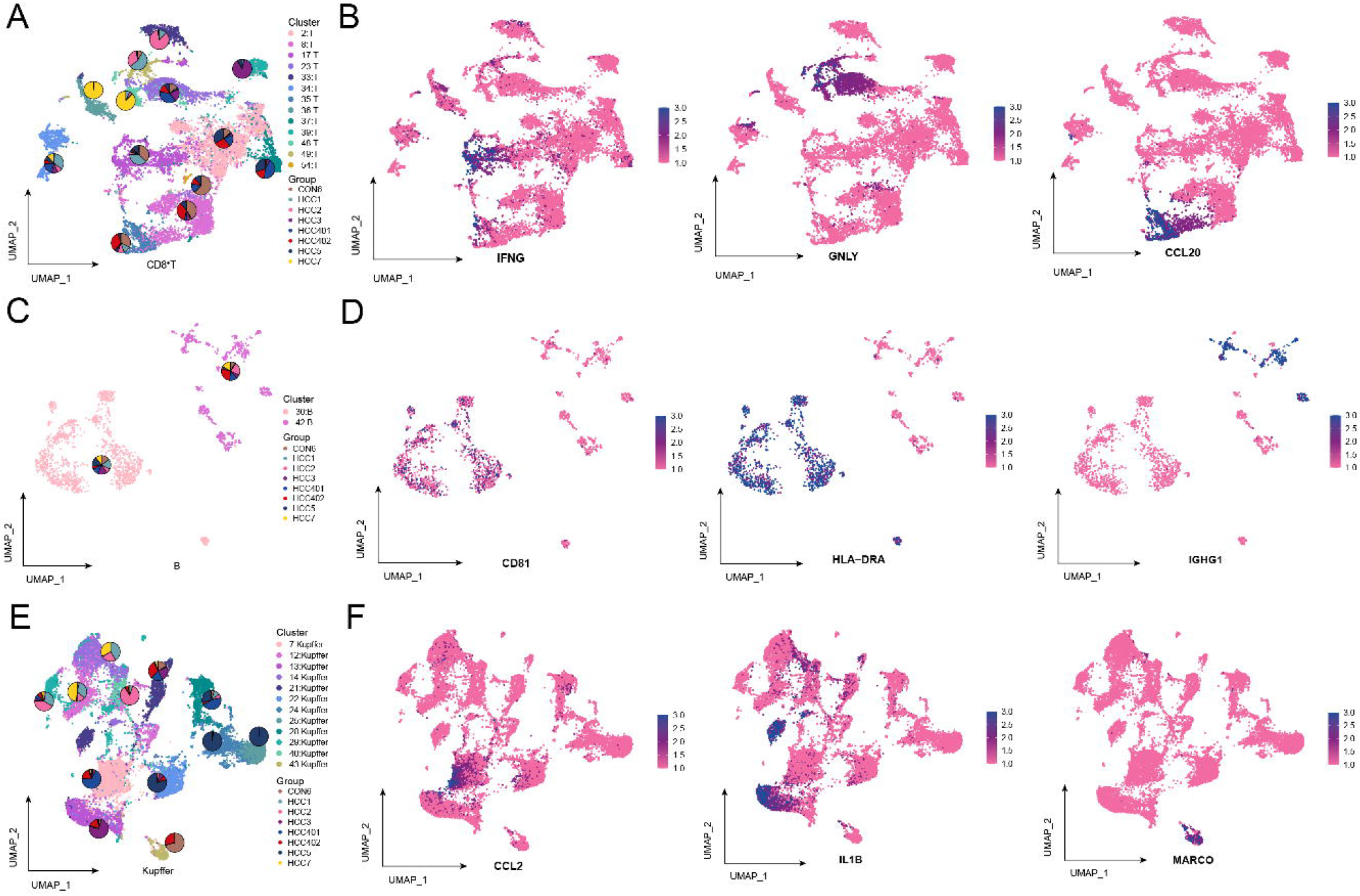
Single-cell mapping of specific immune cells in hepatocellular carcinoma (HCC). A. 13 subsets of CD8+ T cells. The proportion of HCC patient and control donor cells is represented by a pie chart. B. Single-cell atlas mapping immune-related gene expression in CD8+ T cells. C. 2 subsets of B cells. D. The single-cell atlas mapping immune-related gene expression in B cells. The proportion of HCC patient and control donor cells is represented by a pie chart. 12 subpopulations of E. Kupffer cells. The proportion of HCC patient and control donor cells is represented by a pie chart. F. The single-cell atlas mapping immune-related gene expression in Kupffer cells.

### Clonal evolutionary pattern of HCC cells

To understand the role of HCC cells in the evolution of disease progression, we first extracted HCC cells for reclustering and obtained 15 HCC cell subpopulations (Figure 5A). CNV estimation was performed on HCC cell subsets, and the CNV of HCC lesions and liver cirrhosis lesions (HCC401 and HCC402) in the same patient were essentially the same (Figure 5B). Pseudotime trajectories of hepatocyte development showing the development of malignant subclones of HCC revealed that the HCC_7 subpopulation was in early development while the HCC_3 subpopulation was in late development (Figure 5C). In addition, the gene regulatory network with transcription factors (TFs) as the fulcrum was organized into four modules (Figure 5E), such as TP53, HOXA7, FOXN3 and PPARG, to regulate the RNA transcription rate of the HCC malignant cell subpopulation (Figure 5D). In conclusion, these results suggest that multiple malignant cell subsets exist in HCC patients, which may develop from some liver disease (cirrhosis) cells, and that the specific gene expression of HCC malignant cell subsets is regulated by some transcription factors, which in turn guide cell fate. Selection promotes the transformation and differentiation of the core state and ultimately mediates the formation of a range of clonal phenotypes.

**Figure 5:**
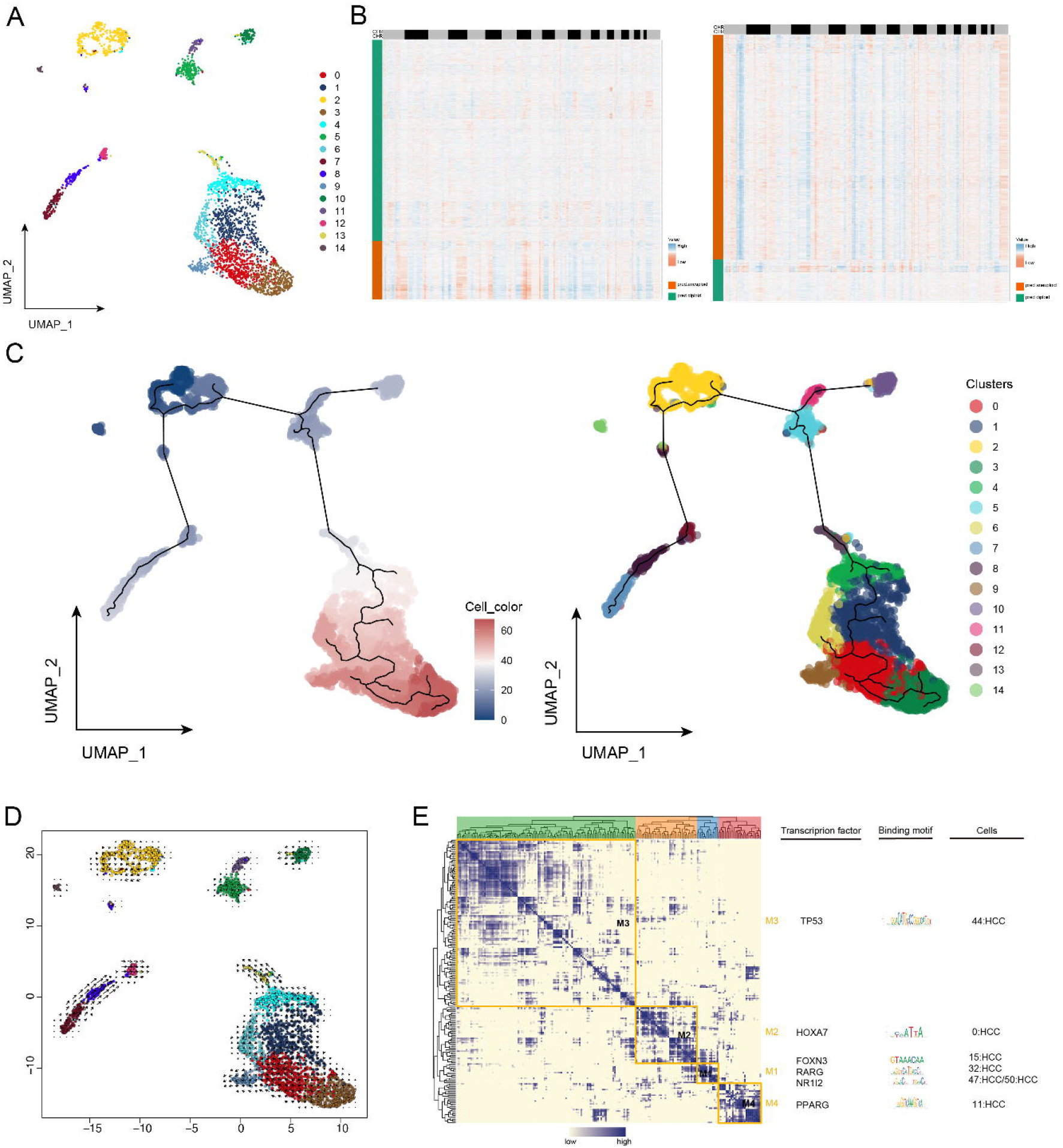
Exploration of the clonal evolution pattern of malignant cells in hepatocellular carcinoma (HCC) cells. A. The single-cell atlas of malignant subclones of HCC. B. Chromosomal heatmap showing copy number variation estimates for two lesions, HCC401 (left) and HCC402 (right). C. Pseudochronological developmental and differentiation trajectories of hepatocytes. D. Mapping of RNA rates of HCC malignant subclones in a single-cell atlas. E. Co-expression modules of transcription factors in HCC malignant subclones of HCC patients. Left: The regulator module is identified from the regulator’s connection specificity index matrix. middle. The representative transcription factors and their binding motifs in modules. Right panel: The association of modules with malignant subclones.

### Early recurrence ecotone of HCC

The challenge of a high relapse rate after radical treatment of HCC remains. During the study, one patient (HCC2) was found to have an early postoperative relapse with a spatial single-cell and multi-omics molecular pattern worthy of investigation. Therefore, the single-cell landscape of this patient after relapse was explored (Figure 6A), and 54 clusters and 14 cell types were obtained. CNV was observed at the genomic level in patients with early recurrent HCC (Figure 6B), and somatic mutation spectrum (Figure 6C) and fusion gene patterns (Figure 6D) in patients with early recurrent HCC. More CNVs were present in patients with recurrent HCC than in those without recurrence. Thus, HCC cell heterogeneity and high CNVs may contribute to recurrence.

**Figure 6:**
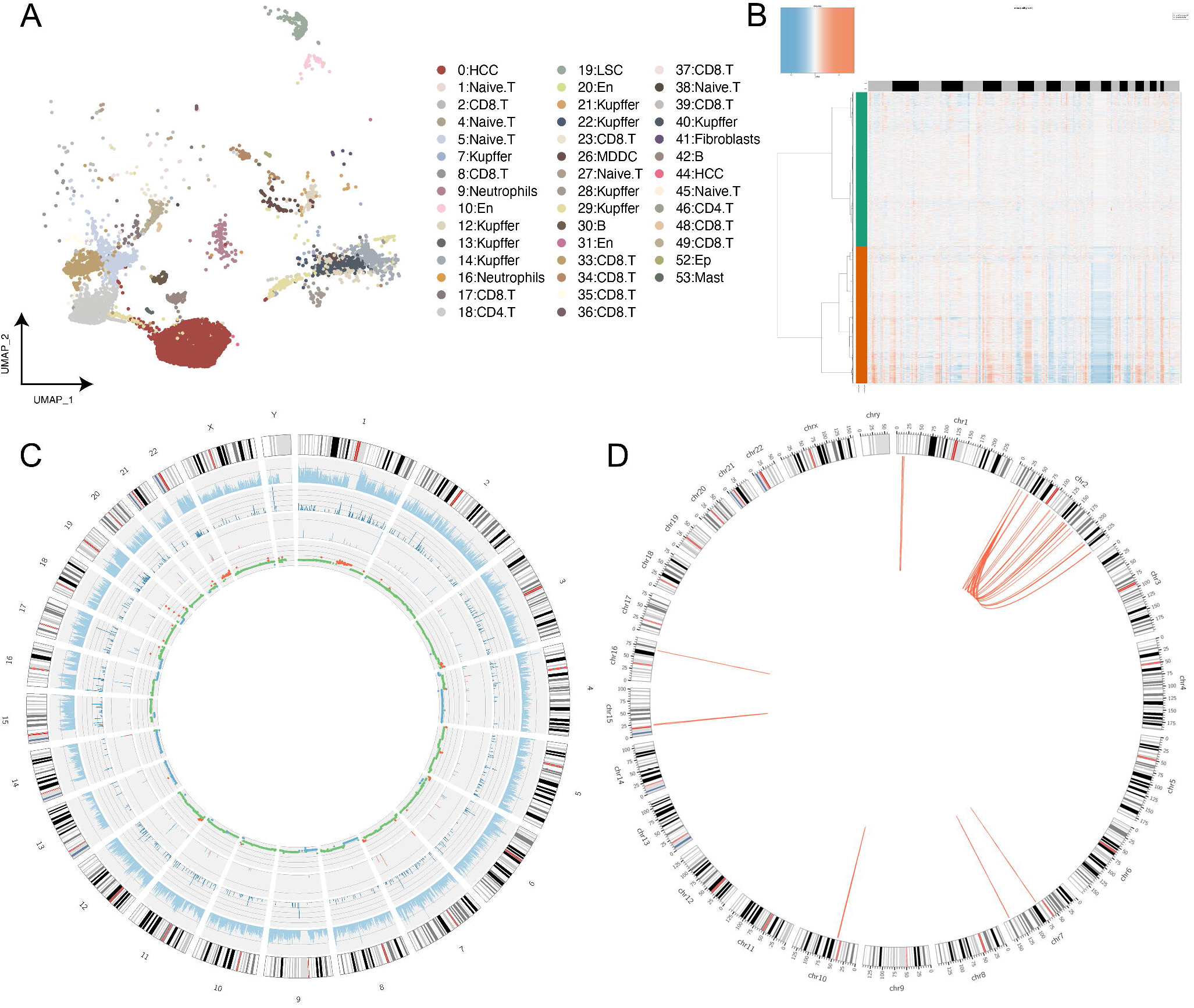
Early recurrence niche of hepatocellular carcinoma (HCC). A. The single-cell atlas maps the single-cell landscape of patients with early stage recurrent liver cancer. B. The chromosome heatmap and chromosome bar graph showing the spectrum of copy number variation in patients with early recurrent HCC. C. The chromosomal circPlot showing the spectrum of somatic mutations in patients with early stage recurrent HCC. D. The chromosomal circPlot connection diagram showing the fusion gene pattern in patients with early recurrent HCC.

## Discussion

So far, novel small molecular targeted and immunotherapy regimens greatly improved the multidisciplinary treatment of HCC ^[35]^. However, rare promising results have been reported in phase III clinical trials, the response rates remains suboptimal, owing to immune escape events mediated by tumor molecular heterogeneity ^[36]^. In the current study we conducted single-cell technology, ST technology, and ONT to generate an innovative, direct and in depth depiction of the ecological niche of HCC tumor heterogeneity and its clonal evolutionary pattern.

Our results demonstrated the existence of multiple subpopulations of malignant cells in patients with HCC, suggesting that the intra-tumor heterogeneity in HCC stems primarily from the diversity of tumor cells. However, the fact that tumors are not a single clonal genome; rather, multiple distinct subclones that evolve from one or more origins during the disease cycle of HCC development, thereby mediating intra-tumor heterogeneity. Therefore, we performed reclustering and CNV estimation of the HCC cells. The results showed that the CNA of the two lesions (HCC lesions and cirrhosis lesions) in the same HCC patient were basically the same, suggesting that the two lesions may have a common origin, and the lesion cells gradually differentiated into HCC cells. GRN analysis showed that malignant cell subpopulations of HCC are regulated by multiple TFs. Among them, the link between HCC progression and TP53 mutations has been demonstrated ^[37]^, and the linkage between HOXA7, FOXN3, NR1I2, and PPARG has also been well documented ^[38-41]^. In contrast, the association of RARG with HCC has rarely been reported in previous studies, and its dysregulation was found to be associated with acute promyelocytic leukemia (APL) ^[42]^. In the present study, we found that RARG is involved in the regulation of malignant cell subpopulations in HCC and may be a novel potential therapeutic target for HCC. Taken together, these findings led us to hypothesize that malignant cells of common origin with very few differential CNAs and expression patterns gradually evolve to form distinct foci and are mainly regulated by expression patterns, thus, mediating intertumor heterogeneity.

Metastasis in HCC is closely associated with angiogenesis^[43]^ and that the remarkable plasticity of endothelial cells contributes to pathological angiogenesis ^[44]^ is increasingly certain. Therefore, we explored the endothelial cell subpopulation in HCC. In the present study, the endothelial cell subpopulation was involved in the resistance pathway to epidermal growth factor receptor tyrosine kinase inhibitors, suggesting that patients with HCC have developed resistance to these drugs. Subsequent GRN analysis showed that endothelial cells were regulated by SREBF1, ZNF585A, and HOXB5, and the remarkable expression of exosome marker genes was observed in both malignant and endothelial cell subpopulations. Among them, SREBF1 appears to be associated with cancer ^[45]^ but is rarely reported in HCC, whereas HOXB5 promotes HCC metastasis by targeting FGFR4 and CXCL1 ^[46]^. HCC-derived exosomes are the main drivers of pre-metastatic ecotone formation and can mediate the metastasis of tumor cells to specific organs ^[47, 48]^. Thus, we tentatively inferred that angiogenic ecotone genes mediate the communication between HCC subpopulations and endothelial cells through exosomes, contributing to the pre-metastatic angiogenic ecotone of HCC.

Next, we explored the ecological niche of immune cell subpopulations in HCC and found that Kupffer cells are widely present in patients with HCC. Kupffer cells act as resident hepatic macrophages, both as effector cells that destroy hepatocytes and antigen presenting cells during hepatic viral infection ^[49]^. Interestingly, we found that CCL2, IL1B, and MARCO were remarkably expressed in the Kupffer cell subpopulation. Previous studies have confirmed that higher MARCO expression is associated with poor prognosis in a variety of cancers ^[50-52]^. More importantly, preclinical studies have shown that anti-MARCO antibodies inhibit tumor growth and metastasis in 4T1 breast cancer and B16 melanoma mouse models ^[53]^. This suggests that in HCC, high MARCO expression may be involved in HCC immunomodulation by regulating Kupffer cells and may be an interesting therapeutic target for HCC. In addition, we explored the ecological niche of patients with early relapse of HCC, considering that relapse of HCC is the ultimate cause of treatment failure and death. More copy number variants were present in patients with recurrent HCC than in those without recurrence, and these mutated genes may be HCC early recurrence ecotype genes.

This study remains several limitations. We included only six patients with HCC and one control in our study; therefore, these results need to be further validated in a larger sample. In addition, our results were derived using scientific bioinformatics analysis methods, which were not demonstrated in experimental or clinical practice. As the establishment of animal models for such purposes is difficult, clinical trials may provide valuable information.

In summary, we performed a multi-omics approach to conduct a comprehensive study of tumor heterogeneity and its clonal evolutionary patterns to reveal heterogeneity-mediated progression and recurrence of HCC in tumor cells under the tumor microenvironment. These findings described the great potential prognostic and therapeutic targets for HCC with decision-making value in the clinical practice.

## Supporting information

Demographic and clinical data are presented in Table S1

## Figure legend

**Supplementary Table 1. Patient Clinical Information**.

